# Assessing respiratory rates and signs of air hunger using the PneumoRator® sensor

**DOI:** 10.64898/2026.09.21.26363466

**Authors:** JJ. Hudson-Colby, H. Baston, M. Harding, S. Brider, F. Yates, M. Shaban, N. White, A. Lewis

## Abstract

**Objectives:** To investigate whether the PneumoRator wearable sensor could accurately assess Respiratory Rate (RR) and signs of air hunger (SAH) compared with clinician-observed assessment in adults with and without breathing pattern disorder (BrPD).

**Design:** A pilot cross-sectional observational study.

**Setting:** A research laboratory on a university campus.

**Participants:** Healthy adults aged 18-40, who were not pregnant, no smokers or history of cardiovascular, respiratory or neurological illness. Thirty consented to the study, Twenty-nine had full data for analysis.

**Interventions:** Participants wore the PneumoRator chest-worn sensor continuously for 10 minutes while seated at rest.

**Main outcome measures:** Respiratory Rate was compared between PneumoRator-derived measurements and randomised 1-minute observer manual counts. Clinician-observed SAH and BrPD were assessed using the Breathing Pattern Assessment Tool (BPAT). Participants also completed the Nijmegen Questionnaire. PneumoRator signals were analysed to identify large-volume breaths as potential SAH events.

**Results:** Thirty participants were recruited. Twelve participants met BPAT criteria for BrPD and four met Nijmegen Questionnaire criteria. There was no significant difference between manual and PneumoRator RR measurements (p=0.89). No significant difference was observed between methods for SAH detection (p=0.17). Participants with clinician-observed SAH demonstrated significantly higher PneumoRator signal amplitudes than those without SAH (p=0.035).

**Conclusion:** The PneumoRator produced RR measurements comparable to clinician assessment and demonstrated potential for identifying signs of air hunger. Findings from this pilot study indicate that the PneumoRator device shows promise as a tool for continuous RR monitoring and as an adjunct to BrPD assessment. Larger clinical validation studies are now warranted.

## Background

Respiratory Rate (RR) is an important predictor of clinical deterioration [1,2,3]. Regardless of its proven significance, RR measurements are viewed as burdensome [4], impeded by interruptions, time and equipment constraints, and clinical ignorance [2,5,6].

The gold standard measurement technique for RR is contested, with evidence split between 1-minute manual counts [3,7] and capnography [8,9]. Gold standard techniques enable prompt and accurate identification of deterioration features that may be missed by other vital signs [10]. Gold standard disputes may arise from measurement limitations and errors reported in the literature. Despite the ease of manual measurements, Palmer *et al*. [6] summarised that they regularly underestimate RR. As such, variations in RR can go undetected [4, 11]. Conversely, capnography provides accurate, prolonged measures, but functional attachments impede patient comfort, practicality, and bias RR values [12].

Exploration of novel technologies for RR measurement is ongoing, although current evidence remains confined to singular, validation studies [1,11,12]. Among emerging options, contact triaxial accelerometers, such as the PneumoRator device, offer lightweight, discreet measurement of chest wall movement through capacitance sensors [13]. Custom algorithms attenuate motion artifacts that may impact accelerometer-based technologies [12], allowing normal activities free of restrictive piezoelectric belts or capnography facial attachments. Although the PneumoRator satisfies proposed requirements for seamless integration into clinical practice [8,14], it lacks ecological validity and clinical validation.

### Breathing Pattern Disorder and Measurement

Breathing pattern disorder (BrPD) is an alteration of regular breathing, caused by functional or structural irregularities [15, 16, 17]. BrPD can be a standalone diagnosis, or it can be part of a multimorbid presentation such as with asthma [15] or other health conditions [16, 18].

The most common diagnostic tool is the Breathing Pattern Assessment Tool (BPAT): a partially objective, multicomponent observational tool [18,19,20,21]. Clinically assessed scores of ≥4 are highly sensitive and specific for indicating BrPD [20]. An alternative measure is the Nijmegen Questionnaire (NQ). This is a self-reported measure that investigates functional respiratory complaints [15,18] with a diagnostic cut off of 19/64 or 23/64 depending on the coexisting diagnosis of Asthma [16,22,23]. The detection of significant changes in breathing pattern, such as sighing and deep breathing or yawning, is integral to accurate diagnosis and prognosis [24]. Severs, Vlemincx and Ramirez [24] define sighing and yawning as “signs of air hunger” (SAH), which forms part of the BPAT used in the diagnosis of BrPD. Accurately quantifying the frequency and characteristics of yawning or sighing using non-invasive technology has been complex. Firstly, the current objective definition for Air Hunger is derived from spirometry studies: Tidal volume (Vt) >2x the average Vt [24].

RR accuracy significantly improves through minimal increases in measurement length [6], and so may improve over prolonged periods. Furthermore, RR is currently the least frequently automated vital sign, highlighting the absence of a clinical technology for RR monitoring [11, 25]. Using the PneumoRator for sustained RR measurement may mitigate the cumulative demand of vital sign monitoring on healthcare professionals [4], releasing them to perform other caring duties. Furthermore, having an objective measurement of air-hunger recorded over a minute would enhance current assessment methods for breathing pattern disorder and could offer tool for use in treatment and monitoring.

### Objective

This study aimed to compare the PneumoRator-driven continuous assessment of RR and SAH with clinician-observed assessment.

## Methods

### Recruitment

Participants were recruited using convenience sampling on a university campus, using posters with QR codes linked to a Microsoft Forms sheet to register interest or request further information. Eligible volunteers were emailed, attaching the Participant Information Sheet (PIS) and a choice of available dates and times to participate. The sample size was determined by the timeframe available and research laboratory access during the undergraduate dissertation data collection timelines.

### Eligibility

#### Inclusion Criteria

Healthy individuals who could provide informed consent, with no known history of respiratory, cardiovascular, or neurological disorders, aged 18-40.

#### Exclusion Criteria

Presence of any acute illness at the time of assessment, smoking or vaping within the past 12 months, pregnant, and anyone with an inability to remain seated calmly for 10 minutes.

### Procedure

The data collection sessions were conducted in a university research laboratory, using a high-backed chair and laptop on a table. At least three researchers were present at each data collection session. A YouTube video of London road traffic [26] was available for the participants who wished it. This aimed to distract the participants from actively monitoring their breathing by mimicking daily experiences, while avoiding altered RR from strong emotional responses from alternative options.

Participants took part in one assessment session lasting approximately 15 minutes, five minutes for device calibration and 10 minutes for data collection.

The PneumoRator device (Southampton, UK) was attached to the participant using a silicone-based double-sided adhesive, to the top left of their chest, medial third of clavicle, three finger widths below clavicle.

The primary outcome measures were RR and SAH (collected as part of the BPAT) and the RR and SAH during 10 minutes of PneumoRator data. The BPAT was conducted at a randomised minute over the 10 minutes. The randomisation list was generated online [27]. One researcher completed the BPAT providing the primary score with a secondary researcher also performing the BPAT to corroborate results. A score of at least 4 was used to signify BrPD.

The PneumoRator data were exported into MATLAB, where it was converted into capacitance graphs. These graphs were further filtered, enabling both visual and automated RR analysis and the detection of visual fluctuations.

Any participant exhibiting results indicative of BrPD, a BPAT score of 4+ [19], received an email after their visit signposting them to relevant medical advice.

### Statistical Analysis

Statistical analysis was conducted using SPSS Statistics Version 29 Windows (v29.0.1.0). Normality testing revealed PneumoRator RR was non-parametric. Manual and observer RRs were parametric but near the threshold for being non-parametric. A Paired T-Test was used to assess accuracy and inter-rater reliability of main and observer manual RR. A Wilcoxon signed ranks test was conducted between the main manual and PneumoRator RR measurements. A Spearman’s Rho was used for correlation analysis of SAH events between Observer and PneumoRator events. Data are presented with both Medians and interquartile ranges (IQR), and the mean and standard deviation depending on their distribution. Confidence intervals (CI) were reported at 95%. Statistical significance was determined as p<0.05.

PneumoRator data were analysed using the filtered data output in MATLAB. Breaths within the randomly selected minute segment with visibly greater amplitude/duration than contiguous breaths were manually identified as large-volume events.

For each selected minute, the total area under the curve (AUC) was calculated and divided by the PneumoRator-derived respiratory rate to obtain an average AUC per breath for that minute. The AUC of each large-volume breath was compared to this average value. Breaths with an AUC >2 times the average AUC for the selected minute were classified as SAH, in line with previously reported spirometric thresholds [24]. A Wilcoxon signed-rank test was used to compare the number of air hunger events identified by the PneumoRator with those identified by clinician observation. A Mann-Whitney U test was performed to assess differences in AUC values between participants with and without observed air hunger.

AUC analyses were conducted by a researcher that did not perform the BPAT assessment.

### Ethics

Ethics was obtained from the University of Southampton Faculty Ethics Committee via the ERGO II (Ethics and Research Governance Online) site, ethics number 106871. Research was conducted according to the principles of the Declaration of Helsinki.

## Results

Data were collected between October 2025 and January 2026. The pilot study ended once the sample size target was reached. Data were available for 29 participants who had full PneumoRator and manual RR recordings. Please see participant screening and exclusion in figure 1 below:

**Figure 1:**
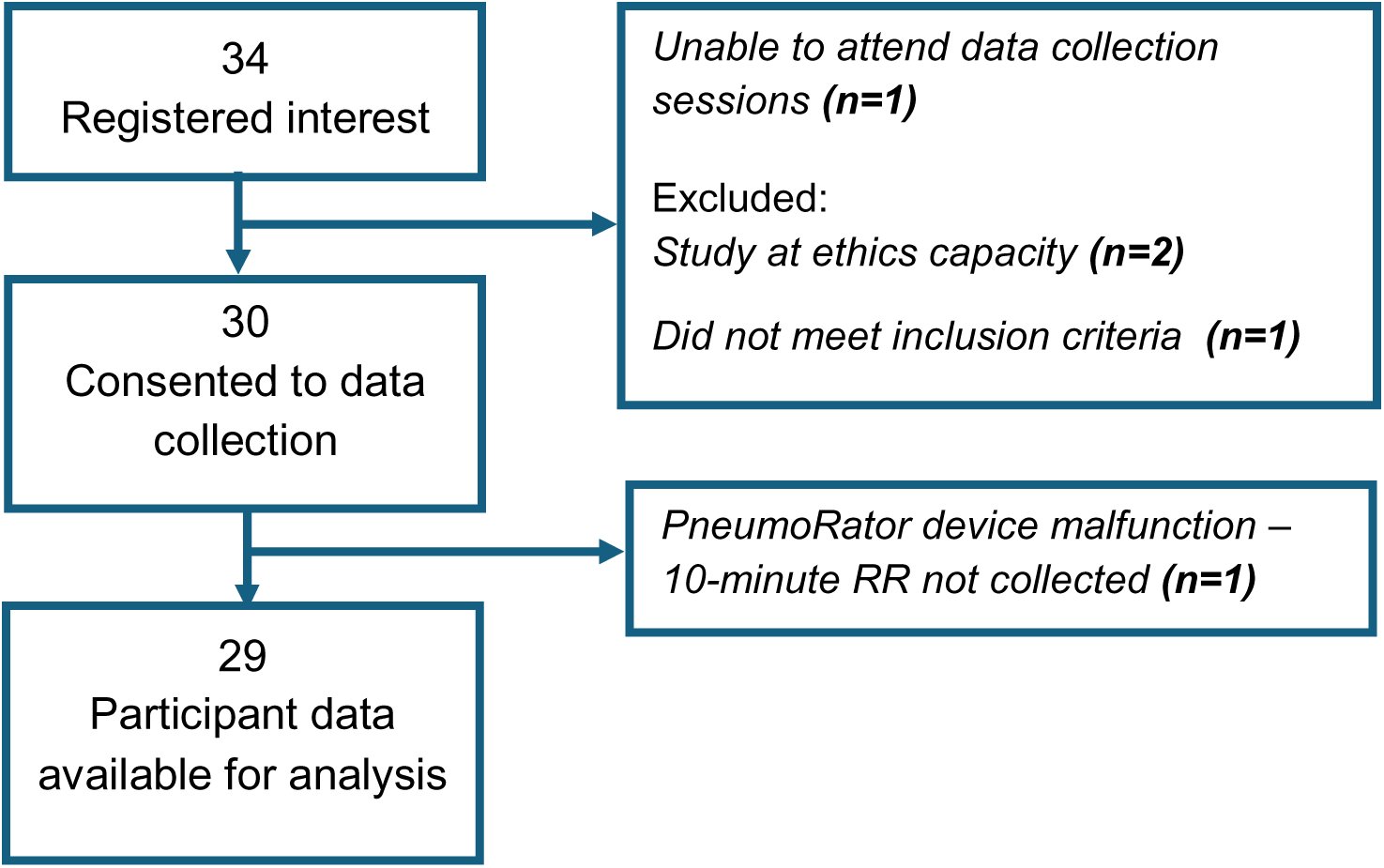
Participant recruitment flow diagram.

**Table 1:** Participant demographics.

|  | Mean (SD) |
| --- | --- |
| Height (cm) | 171.7 (10.2) |
| Weight (Kg) | 72.4 (11.8) |
| BMI | 24.6 (3.2) |
| Age (Years) | 21.2 (3.4) |
|  | N (%) |
| Male | 13 (44.8%) |
| Female | 16 (55.2%) |
|  | Mean (SD) |
| BPAT Score | 3.1 (1.6) |
| NQ Score | 10.3 (7.9) |
| SAH Events (Manual Recordings) | 0.6 (0.7) |

No significant differences were observed between assessors of the BPAT scores taken simultaneously (*t*= 0.069, *p*= 0.65; 95% CI, -0.24 to 0.37). This increases confidence in categorisation of those individuals with and without BrPD from a clinician’s observation.

The median and IQR of manual and PneumoRator RR are displayed in Table 2 and Figure 2. Results of the Wilcoxon signed ranks analysis showed no significant difference between the two methods (*w*= -0.141, *p*= 0.89; 95% CI= -2.28 to 1.5). The median difference between variables (Manual minus PneumoRator) was -0.67 (95% CI, -1.89 to 1.78), falling within the minimally clinically important difference of ±2 bpm when measuring RR.

**Figure 2:**
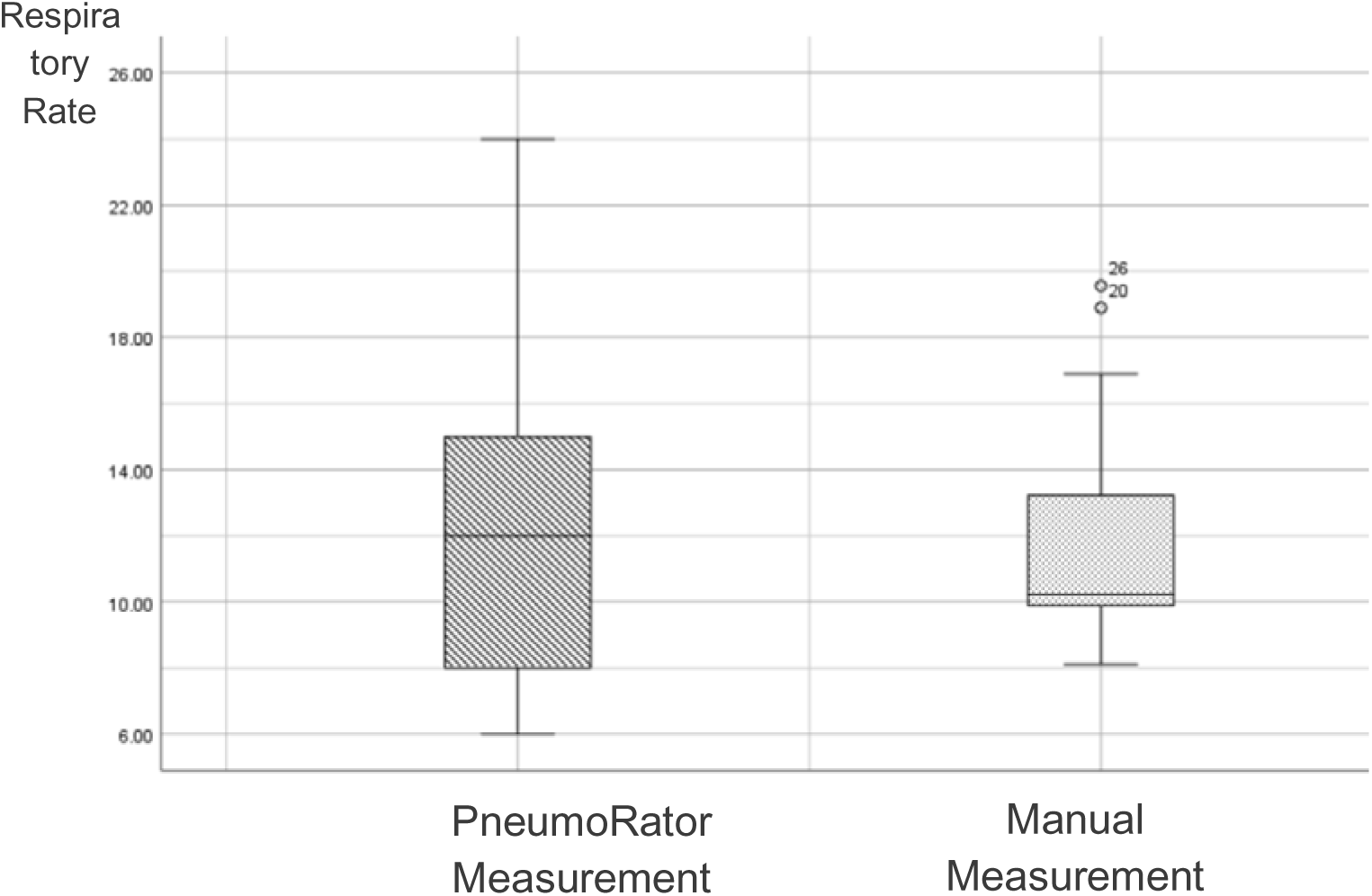
Box and Whisker plot showing the distribution (minimum, 1^st^ quartile, median, 3^rd^ quartile and maximum values) of RR values gathered by the PneumoRator and manual measurements.

**Table 2:**
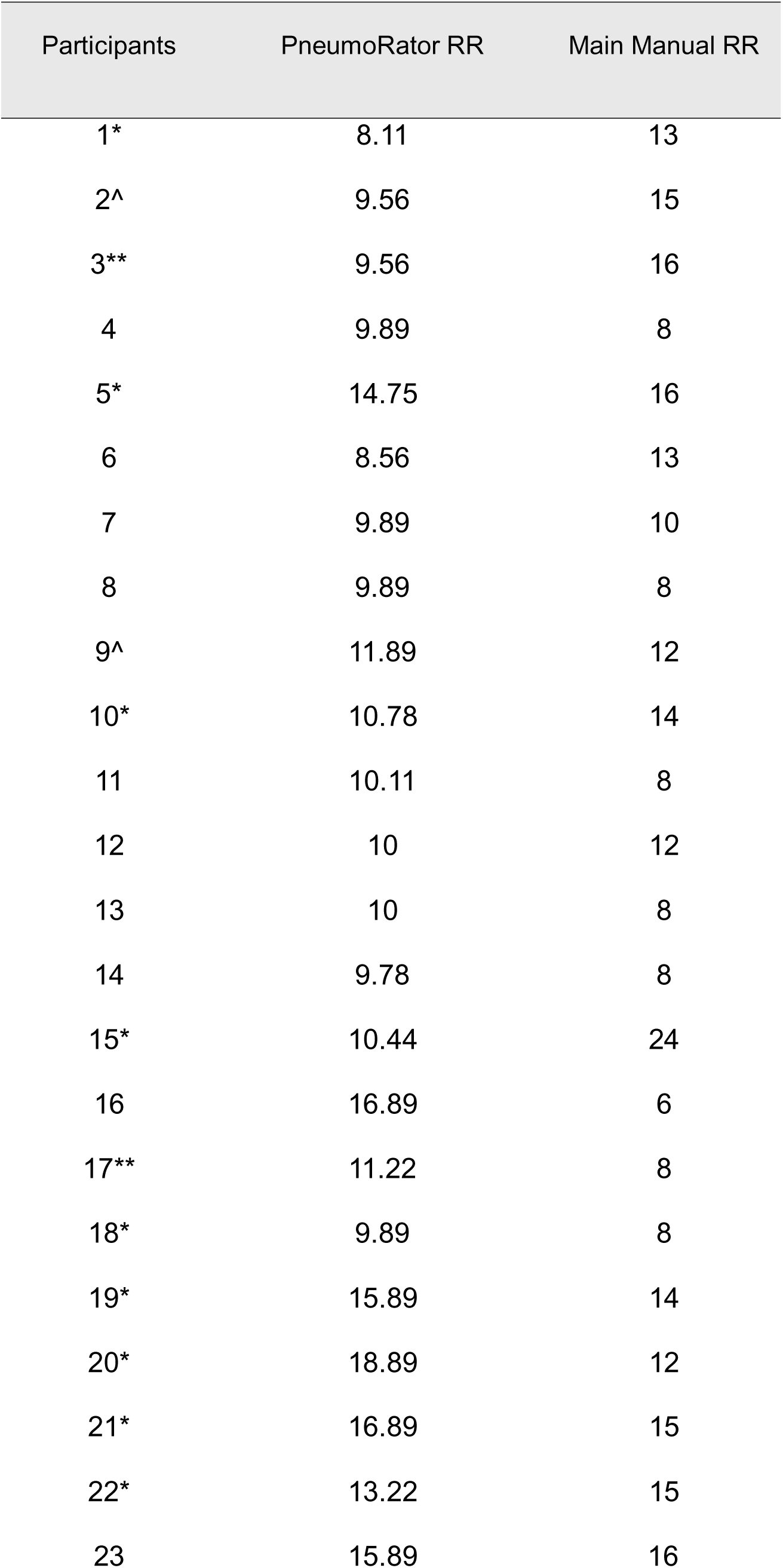
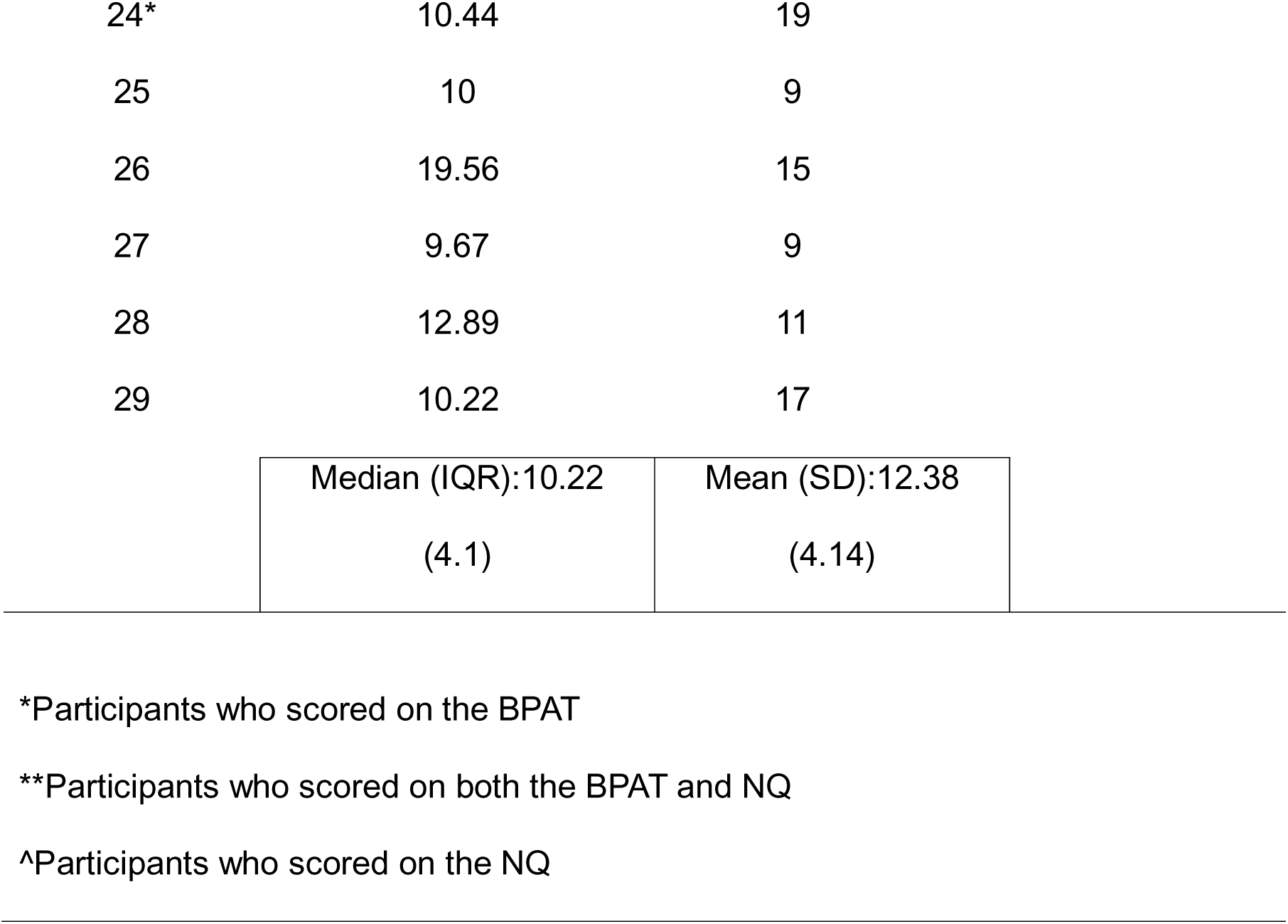
Individual participant RR data obtained using the PneumoRator and manual measurements.

Twelve participants had BrPD according to the BPAT, whilst only four participants had BrPD according to the NQ cut off score of 19, and two had BrPD according to combined measures.

The odds of participants being diagnosed with BrPD using the BPAT and exhibiting increased RR variability from PneumoRator monitoring (SD >4 bpm) were high but insignificant (Odds ratio= 3.2; 95% CI, 0.26 to 40.06, p=0.55).

### Signs of Air Hunger

Nineteen out of 29 (66%) participants were in exact agreement regarding number of air hunger events between BPAT and PneumoRator data. No statistically significant difference was observed between the two assessment methods (Z = -1.387, p = 0.17). A statistically significant, moderate, positive correlation is observed between the methods (ρ = 0.50, p = 0.006). Further per participant data are shown below in table 3.

**Table 3:** Largest PneumoRator AUC values and clinician-observed breathing pattern disorder (BrPD) status according to the BPAT.

| Participants | Largest AUC for selected minute | BrPD according to BPAT |
| --- | --- | --- |
| 1 | 3.50 | Yes |
| 2 | 1.43 | No |
| 3 | 1.11 | No |
| 4 | 1.25 | Yes |
| 5 | 1.67 | No |
| 6 | 1.11 | No |
| 7 | 2.25 | Yes |
| 8 | 1.11 | No |
| 9 | 2.32 | No |
| 10 | 1.85 | No |
| 11 | 1.46 | No |
| 12 | 1.02 | No |
| 13 | 2.63 | No |
| 14 | 1.69 | No |
| 15 | 1.19 | No |
| 16 | 3.93 | Yes |
| 17 | 1.65 | No |
| 18 | 0.87 | Yes |
| 19 | 1.50 | Yes |
| 20 | 1.90 | Yes |
| 21 | 2.60 | Yes |
| 22 | 2.08 | Yes |
| 23 | 1.30 | No |
| 24 | 2.01 | Yes |
| 25 | 1.29 | Yes |
| 26 | 4.02 | Yes |
| 27 | 0.87 | No |
| 28 | 1.42 | No |
| 29 | 1.66 | No |

Participants with clinician-assessed SAH demonstrated statistically significant higher AUC values (mean rank 18.96) compared with those without clinician-assessed Air Hunger (mean rank 12.21), between group difference U= 54.5, Z= -2.105, p= 0.035.

## Discussion

This study aimed to investigate whether the PneumoRator could be used to monitor RR and SAH in adults with and without BrPD.

Our first novel finding is that there were no significant differences between manual and PneumoRator RR counts or counts of SAH in both healthy adults and those with BrPD. Unlike most existing evidence that compares new with established devices [1,3,11], Bawua *et al*. [8] discussed device (impedance pneumography and electrocardiographic-derived respiration) vs clinician measurement. This systematic review highlighted inconsistent findings depending on the analysis method. Results should be interpreted cautiously as statistical similarity may not mean methods are clinically interchangeable.

Our second novel finding is that in those participants with clinically assessed SAH, the AUC values were higher than those without clinically assessed SAH. Despite its clinical significance, the study by Severs, Vlemincx and Ramirez [24] focuses on the physiology of a sigh, a complex breathing phenomenon that occurs spontaneously in normal breathing. The lack of current research on SAH prompted this study to explore this definition and determine whether it could be applied to the PneumoRator. Results suggest that whilst the >2x AUC threshold did not perfectly match clinician counts, participants with *observed* air hunger had significantly higher AUCs. There may need to be further mathematical calibration of this ‘>2x’ threshold to optimise clinical validity in future studies.

These two findings together suggest that the PneumoRator captures breathing events that align with the clinician observation of RR and air hunger. Therefore, the PneumoRator could be used as a valuable tool in clinical practice to continually monitor RR and signs of air hunger to have a more sensitive diagnosis and understanding of a person’s BrPD.

Our third novel finding is that this is the first study recruiting healthy volunteers who then get screened for BrPD using both BPAT and NQ data. Our study shows the discrepancy in prevalence according to these assessment methods away from a respiratory outpatient clinic environment. Literature remains unclear about gold standard diagnostic methods for BrPD, generating uncertainty around accurate prevalence rates and preferred diagnostic tools to be used in clinical practice. The lack of a significant odds ratio between RR variability and BrPD diagnosis further supports the need for holistic diagnosis, albeit this could be a type 2 measurement error. While the PneumoRator may provide the valid identification of RR variability spot check measurements lack [4,11], erratic breathing patterns are only a proportion of all BrPD presentations. This highlights a need for a multidimensional diagnostic tool incorporating continuous RR measurement, objective visual breathing assessment, and patient-reported outcomes. Such a combination would reflect the multi-component nature of BrPD, viewing diagnosis holistically rather than debating the utility of individual measures. This may also mitigate biases and improve diagnostic rigour [15]. The PneumoRator could be used as an effective tool in multi-component BrPD diagnosis. Although a limitation, our data confirm the difficulty in diagnosing BrPD using a single outcome measure.

## Limitations

The primary limitation of this study is the small sample size to infer generalisability with high confidence. However, the sample size is adequate for a pilot study investigating a novel technology in this population.

The research environment may have been unrealistic regarding real-world demands on RR. Abnormalities in RR may be more pronounced in the presence of factors such as stress [28] or exercise [29]. This raises discussion around the ecological and external validity of this study, however BrPD can still be disabling when experienced at rest.

At present, the PneumoRator assessment of air hunger is only semi-automated. It requires an assessor to manually identify large-volume breaths from the visualised data before mathematical calculation can occur. This also introduces the further risk of analyst selection bias, as the mathematical objectivity of the AUC calculation is compromised by a subjective visual pre-filtering stage, decreasing the chance of inter-rater reliability. This, however, does not negate the utility of the PneumoRator, especially as a device used in conjunction with clinician assessment, such as the BPAT.

A further limitation is presented through the quantification of BPAT scores. A RR of under 12 is considered healthy, scoring zero, while RR over 25 contributes two points to overall BPAT score. While evidence agrees with the higher limit showing clinical criticality [7,30], low RRs that are of high tidal volumes, may still lead to hyperventilation and cause BrPD.

## Conclusion

This study compared PneumoRator device RR and SAH measurements over 10 minutes with 1-minute manual measurements and BPAT assessed SAH in adults with and without BrPD. Methodological restrictions limit the generalisability to wider populations, however data here support that further studies in larger samples in clinical environments are now warranted because the PneumoRator shows potential to be used as a tool for respiratory rate and BrPD assessment and monitoring.

## Data Availability

Data sharing may occur on reasonable request.

## Data Sharing Statement

Data sharing may occur on reasonable request.

## Author Contributorship

JJ Hudson-Colby, Adam Lewis, Neil White and Mahdi Shaban conceptualised the study, JJ Hudson-Colby and Adam Lewis led the study and supervised, Holly Baston, Shannon Brider, Melissa Harding and Finlay Yates. Holly Baston, Shannon Brider, Melissa Harding and Finlay Yates, JJ Hudson-Colby and Adam Lewis collected the data. Holly Baston, Shannon Brider, Melissa Harding and Finlay Yates analysed the data. Adam Lewis wrote the first draft of the paper, all other authors revised further drafts. All authors agreed the final draft for submission.

## Acknowledgements

We thank all study participants for their time. Microsoft Copilot AI was used to support the structuring of the abstract according to journal guidelines, which were then further edited by Adam Lewis.

## Contribution of the paper

- There is no commonly-used continuous objective measure of “signs of air hunger” in physiotherapy breathing pattern disorder assessment, and spot check manual assessments of respiratory rate can be inaccurate.
- There is no significant difference in respiratory rate assessment between continuous PneumoRator® chest-worn sensor and manual clinician-observed methods.
- The PneumoRator also identified signs of air hunger that matched clinician observations, with clinically observed signs of air hunger showing larger breathing signals on the PneumoRator.

## References

1. Sato H, Nagano T, Izumi S, et al. Prospective observational study of 2 wearable strain sensors for measuring the respiratory rate. Medicine (Baltimore). 2024 Jul 19;103(29):e38818. doi: 10.1097/MD.0000000000038818. PMID: 39029069; PMCID: PMC11398755.

2. Hamada O, Tsutsumi T, Tsunemitsu A, et al. Improving respiratory rate monitoring in general wards following implementation of a rapid response system: a quality improvement initiative. BMJ Open Qual. 2025 Apr 9;14(2):e003218. doi: 10.1136/bmjoq-2024-003218. PMID: 40210248; PMCID: PMC11987154.

3. Stevens G, Larmuseau M, Damme AV, et al. Feasibility study of the use of a wearable vital sign patch in an intensive care unit setting. J Clin Monit Comput. 2025 Feb;39(1):245–256. doi: 10.1007/s10877-024-01207-5. Epub 2024 Aug 19. PMID: 39158782.

4. Posthuma LM, Downey C, Visscher MJ, et al. Remote wireless vital signs monitoring on the ward for early detection of deteriorating patients: A case series. Int J Nurs Stud. 2020 Apr;104:103515. doi: 10.1016/j.ijnurstu.2019.103515. Epub 2020 Jan 3. PMID: 32105974.

5. Harry ML, Heger AMC, Woehrle TA, et al. Understanding Respiratory Rate Assessment by Emergency Nurses: A Health Care Improvement Project. J Emerg Nurs. 2020 Jul;46(4):488–496. doi: 10.1016/j.jen.2020.03.012. Epub 2020 May 29. PMID: 32482501.

6. Palmer JH, James S, Wadsworth D, et al. How registered nurses are measuring respiratory rates in adult acute care health settings: An integrative review. J Clin Nurs. 2023 Aug;32(15-16):4515–4527. doi: 10.1111/jocn.16522. Epub 2022 Sep 12. PMID: 36097417.

7. Aglen SAS, Simonsen HF, Sjoset TE, et al. Respiratory Rate as a Predictor of Clinical Deterioration and Mortality: A Scoping Review. Acta Anaesthesiol Scand. 2025 Sep;69(8):e70113. doi: 10.1111/aas.70113. PMID: 40828518.

8. Bawua LK, Miaskowski C, Hu X, et al. A review of the literature on the accuracy, strengths, and limitations of visual, thoracic impedance, and electrocardiographic methods used to measure respiratory rate in hospitalized patients. Ann Noninvasive Electrocardiol. 2021 Sep;26(5):e12885. doi: 10.1111/anec.12885. Epub 2021 Aug 18. PMID: 34405488; PMCID: PMC8411767.

9. Kodali, B. Capnography Outside the Operating Rooms. Anesthesiology, 2013 118(1), 192–201. 10.1097/ALN.0b013e318278c8b6.

10. Akel MA, Carey KA, Winslow CJ, et al. Less is more: Detecting clinical deterioration in the hospital with machine learning using only age, heart rate, and respiratory rate. Resuscitation. 2021 Nov;168:6–10. doi: 10.1016/j.resuscitation.2021.08.024. Epub 2021 Aug 23. PMID: 34437996; PMCID: PMC9128300.

11. Drummond GB, Fischer D, Lees M, et al. Classifying signals from a wearable accelerometer device to measure respiratory rate. Eur Respir J Open Research 2021; 7(2). Available at: 10.1183/23120541.00681-2020.

12. Cesareo A, Biffi E, Cuesta-Frau D, et al. A novel acquisition platform for long-term breathing frequency monitoring based on inertial measurement units. Med Biol Eng Comput. 2020 Apr;58(4):785–804. doi: 10.1007/s11517-020-02125-9.

13. Terrington I. Accurately monitoring respiratory rate via a novel wearable device. Hospital Healthcare Europe 2025. Available at: https://hospitalhealthcare.com/clinical/respiratory/accurately-monitoring-respiratory-rate-via-a-novel-wearable-device/

14. Guy EFS, Clifton JA, Knopp JL, et al. Non-Invasive Assessment of Abdominal/Diaphragmatic and Thoracic/Intercostal Spontaneous Breathing Contributions. Sensors (Basel). 2023 Dec 12;23(24):9774. doi: 10.3390/s23249774.

15. Hudson-Colby JJ, Lewis A, Varkonyi-Sepp J, et al. Understanding the impact of breathing pattern disorders in difficult-to-treat asthma. Expert Rev Respir Med. 2024 Oct;18(10):777–788. doi: 10.1080/17476348.2024.2404673.

16. Takeda N, Koya T, Hasegawa T, et al. Prevalence and characteristics of dysfunctional breathing in patients with asthma in the Japanese population. Respir Investig. 2024 Nov;62(6):1015–1020. doi: 10.1016/j.resinv.2024.08.004.

17. Bondarenko J, Burge AT, Bremner J, et al. Nonpharmacological Interventions for Dysfunctional Breathing in Adults: A Systematic Review. J Allergy Clin Immunol Pract. 2025 Aug;13(8):2062–2074. doi: 10.1016/j.jaip.2025.04.053.

18. Reilly CC, Floyd SV, Lee K, et al. Breathlessness and dysfunctional breathing in patients with postural orthostatic tachycardia syndrome (POTS): The impact of a physiotherapy intervention. Auton Neurosci. 2020 Jan;223:102601. doi: 10.1016/j.autneu.2019.102601.

19. Todd S, Walsted ES, Grillo L, et al. Novel assessment tool to detect breathing pattern disorder in patients with refractory asthma. Respirology. 2018 Mar;23(3):284–290. doi: 10.1111/resp.13173.

20. Bondarenko J, Hew M, Button B, et al. Reliability of the breathing pattern assessment tool for in-person or remote assessment in people with asthma. Clin Exp Allergy. 2021 Sep;51(9):1218–1220. doi: 10.1111/cea.13856.

21. Milstein CF, Patel RR, Laurash E, et al. Identification of Breathing Pattern Disorder in Athletes With Exercise-Induced Laryngeal Obstruction: A Novel Assessment Tool. J Voice. 2025 Jul;39(4):1139.e11-1139.e17. doi: 10.1016/j.jvoice.2023.01.006.

22. Reilly CC, Floyd SV, Raniwalla S, et al. The clinical utility of the Breathing Pattern Assessment Tool (BPAT) to identify dysfunctional breathing (DB) in individuals living with postural orthostatic tachycardia syndrome (POTS). Auton Neurosci. 2023 Sep;248:103104. doi: 10.1016/j.autneu.2023.103104. Epub 2023 Jun 17. PMID: 37393657.

23. Van Dixhoorn, J. and Folgering,H. The Nijmegen Questionnaire is useful to quantify and assess the normality of subjective sensations. ERJ Open Res 2015 1(1): 00001–2015 doi: 10.1183/23120541.00001-2015

23. Boulding, R., Stacey, R., Niven, R. et al. Dysfunctional breathing: A review of the literature and proposal for classification. European Respiratory Review, 2016 25(141), pp.287–294. doi:10.1183/16000617.0088-2015.

24. Severs LJ, Vlemincx E, Ramirez JM. The psychophysiology of the sigh: I: The sigh from the physiological perspective. Biol Psychol. 2022 Apr;170:108313. doi: 10.1016/j.biopsycho.2022.108313. Epub 2022 Mar 11. PMID: 35288214;

25. Considine J, Casey P, Omonaiye O, et al. Importance of specific vital signs in nurses’ recognition and response to deteriorating patients: A scoping review. J Clin Nurs. 2024 Jul;33(7):2544–2561. doi: 10.1111/jocn.17099. Epub 2024 Mar 7.

27. Experiencing London. London’s City Ambience: ASMR Traffic Soundscape & 4K HDR Video - City Sounds & Car Sounds Effect! [YouTube]. 2023. Available from: https://www.youtube.com/watch?v=sXJLT3kYdhk

28. RANDOM.ORG. List Randomizer [Internet]. Available from: https://www.random.org/lists/

28. Knöpfel G, Baty F, Uhl F, et al. Quantification of breathing irregularity for the diagnosis of dysfunctional breathing using proportional tidal volume variation: a cross-sectional, retrospective real-world study. BMJ Open. 2024 Jun 16;14(6):e083401. doi: 10.1136/bmjopen-2023-083401.

29. Ionescu MF, Mani-Babu S, Degani-Costa LH, et al. Cardiopulmonary Exercise Testing in the Assessment of Dysfunctional Breathing. Front. Physiol. 2021 11:620955. doi: 10.3389/fphys.2020.620955 11:620955.

30. Candel BG, Duijzer R, Gaakeer MI, et al. The association between vital signs and clinical outcomes in emergency department patients of different age categories. Emerg Med J. 2022 Dec;39(12):903–911. doi: 10.1136/emermed-2020-210628.

